# Mortality of Drug-resistant Tuberculosis in High-burden Countries: Comparison of Routine Drug Susceptibility Testing with Whole-genome Sequencing

**DOI:** 10.1101/2020.08.03.20167213

**Authors:** Martina L. Reichmuth, Kathrin Zürcher, Marie Ballif, Chloé Loiseau, Sonia Borrell, Miriam Reihnhard, Veronika Skrivankova, Rico Hömke, Peter Sander, Anchalee Avihingsanon, Alash’le G Abimiku, Olivier Marcy, Jimena Collantes, E. Jane Carter, Robert J. Wilkinson, Helen Cox, Marcel Yotebieng, Robin Huebner, Lukas Fenner, Erik C. Böttger, Sebastien Gagneux, Matthias Egger, on behalf of the International epidemiology Databases to Evaluate AIDS (IeDEA)

## Abstract

**Background:** Drug-resistant *Mycobacterium tuberculosis* (*Mtb*) strains threaten tuberculosis (TB) control. We compared data on drug resistance obtained at clinics in seven high TB burden countries during routine care with whole-genome sequencing (WGS) carried out centrally.

**Methods:** We collected pulmonary *Mtb* isolates and clinical data from adult TB patients in Africa, Latin America, and Asia, stratified by HIV status and drug resistance, from 2013 to 2016. Participating sites performed drug susceptibility testing (DST) locally, using routinely available methods. WGS was done using Illumina HiSeq 2500 at laboratories in the USA and Switzerland. We used TBprofiler to analyse the genomes. We used multivariable logistic regression adjusted for sex, age, HIV-status, history of TB, sputum positivity, and *Mtb*-lineage to analyse mortality.

**Findings:** We included 582 TB patients. The median age was 32 years (interquartile range: 27-43 years), 225 (39%) were female, and 247 (42%) were HIV-positive. Based on WGS, 339 (58%) isolates were pan-susceptible, 35 (6%) monoresistant, 146 (25%) multidrug-resistant, and 24 (4%) pre-/ extensively drug-resistant (pre-XDR/XDR-TB). The local DST results were discordant compared to WGS results in 130/582 (22%) of patients. All testing methods identified isoniazid and rifampicin resistance with relatively high agreement (kappa 0.69 for isoniazid and 0.88 rifampicin). Resistance to ethambutol, pyrazinamide, and second-line drugs was rarely tested locally. Of 576 patients with known treatment, 86 (15%) patients received inadequate treatment according to WGS results and the World Health Organization treatment guidelines. The analysis of mortality was based on 530 patients; 63 patients (12%) died and 77 patients (15%) received inadequate treatment. Mortality ranged from 6% in patients with pan-susceptible *Mtb* (18/310) to 39% in patients with pre-XDR/XDR-TB (9/23). The adjusted odds ratio for mortality was 4.82 (95% CI 2.43-9.44) for under-treatment and 0.52 (95% CI 0.03-2.73) for over-treatment.

**Interpretation:** In seven high-burden TB countries, we observed discrepancies between drug resistance patterns from local DST and WGS, which resulted in inadequate treatment and higher mortality. WGS can provide accurate and detailed drug resistance information, which is required to improve the outcomes of drug-resistant TB in high burden settings. Our results support the WHO’s call for point-of-care tests based on WGS.

## Introduction

Tuberculosis (TB) is caused by bacteria of the *Mycobacterium tuberculosis* complex (MTBC) and the leading cause of death by a single infectious agent worldwide. In 2018, ten million people were estimated to have developed active TB, of whom 9% were people living with HIV (PLWH). In the same year, around 1.5 million people died from TB, including 251,000 PLWH.^1^ TB accounts for approximately 40% of HIV/AIDS-related adult deaths, and half of these remain undiagnosed.^2^

The emergence of drug-resistant *Mycobacterium tuberculosis* (*Mtb*) strains threatens TB control. In 2018, 3% of new TB cases worldwide were estimated to have multidrug-resistant TB (MDR-TB), and 18% of previously treated cases had MDR-TB.^1^ PLWH are at greater risk of acquiring MDR-TB than HIV-negative people.^3^ Further, treatment outcomes in PLWH co-infected with MDR-TB are worse than among HIV-negative MDR-TB patients.^3^ Pre-extensively and extensively drug-resistant TB (pre-XDR-TB and XDR-TB) pose additional challenges for treatment and control of the disease.^4^ Strategies to control and prevent drug-resistant TB include surveillance, drug susceptibility testing (DST), as well as ensuring the completion of an adequate treatment regimen. However, the treatment of drug-resistant TB is challenging particularly in low- and middle-income countries, due to limited access to DST and effective second-line anti-TB drugs.^1^

The present study is part of a research program on drug-resistant TB of the International Epidemiology Databases to Evaluate AIDS (IeDEA).^5^ In a previous analysis, we compared the results of drug susceptibility testing (DST) performed in high TB burden countries in sub-Saharan Africa, Asia and Latin America with standardized phenotypic DST results obtained at the Swiss National Center for Mycobacteria.^6^ We found that the accuracy of the DST performed at participating sites was moderate and that inadequate DST was associated with increased mortality. The Swiss reference laboratory tested drug resistance to six drugs only: isoniazid (INH), rifampicin (RIF), pyrazinamide (PZA), ethambutol (EMB), amikacin (AMK) and moxifloxacin (MFX). Therefore, other resistances could have been missed, including to streptomycin (SM), kanamycin (KM), ethionamide (ETH), levofloxacin (LFX), or newer drugs.

Whole-genome sequencing (WGS) can simultaneously provide information on resistance to first- and second-line drugs, for which drug-resistance-conferring mutations are known. WGS has the potential to overcome many of the limitations of conventional DST with higher throughput.^7^ We and others showed that drug susceptibility predicted from *Mtb* genomes correlates with phenotypic DST.^8,9^ The World Health Organization (WHO) recommends WGS for drug resistance surveillance and is evaluating sequencing technologies for routine DST.^1^ Here, we compared the drug resistance patterns routinely obtained in seven high TB burden countries with the results from WGS, and examined the mortality associated with discordant resistance profiles using WGS as the reference.

## Methods

As described in detail elsewhere,^6^ we recruited patients from 2013 to 2016 at antiretroviral therapy (ART) programmes and TB clinics serving the same catchment areas in Peru, Thailand, Côte d’Ivoire, Democratic Republic of the Congo (DRC), Kenya, Nigeria, and South Africa. In South Africa, we used strain collections held at the University of Cape Town. All patients had bacteriologically confirmed, or clinically diagnosed TB. We included adult patients (≥ 16 years) with pulmonary TB (PTB) or both PTB and extrapulmonary TB. The recruitment of patients was stratified by HIV status and TB drug resistance as defined at local clinics. We collected demographic and clinical characteristics of participants using a standardised questionnaire. *Mtb* isolates were sub-cultured at the recruitment sites.

### Drug resistance testing

Molecular or phenotypic DST was performed at local laboratories according to routine procedures. DNA was extracted from *Mtb* isolates using standard protocols.^10^ Libraries were prepared using the Illumina Nextera XT kit and sequenced on Illumina HiSeq 2500 at the Department of Biosystems Science and Engineering of the Swiss Federal Institute of Technology (ETHZ) in Basel and the Broad Institute in Cambridge, MA, USA. Sequences had 101, 138, or 151 bp paired-end reads (Illumina, San Diego, CA, USA). We screened one *Mtb* isolate per patient for anti-TB drug resistance mutations using the TBprofiler-v2.8.2 pipeline (https://github.com/jodyphelan/TBProfiler).^7,11^ The pipeline aligns reads to the H37Rv reference genome using bowtie2*-v2.3.5*, BWA-*v0.7.17* or minimap2-*v2.16* and calls variants with SAMtools-*v1.9*.^7,12-15^ The variants were then compared to a drug-resistance database (https://github.com/jodyphelan/tbdb; version of 30.01.2020). Single-nucleotide polymorphism (SNPs) and insertion-deletion (Indels) responsible for resistance to 19 anti-TB drugs were identified:^7,11,16^ SM, para-aminosalicylic-acid (PAS), INH, PZA, cycloserine (CS), KM, ETH, EMB, AMK, RIF, capreomycin (CAP), ofloxacin (OFX), ciprofloxacin (CIP), MFX, LFX, linezolid (LZD), bedaquiline (BDQ), clofazimine (CFZ), and delamanid (DLM). A coverage of 10 reads was needed to call a polymorphism. We considered all drug resistance alleles with a variant frequency equal to or higher than 90%.

### Definitions

WHO defines monoresistance as resistance to one of the first-line drugs (INH-, PZA-, EMB-, and RIF).^1,17^ MDR-TB is defined as resistance to both INH and RIF. Pre-XDR-TB is defined as resistance against INH and RIF plus fluoroquinolones (FQLs) or one of the three second-line injectable (INJ) drugs (AMK, CAP, KM). Extensively drug-resistant TB (XDR-TB) is defined as drug-resistance against INH, RIF, FQLs and at least one of the three second-line INJ drugs.

We compared the drug resistance profiles obtained at local sites to those obtained with WGS. Drug resistance profiles were defined as concordant or as discordant according to the resistance categories defined by WHO.^1^ Discordant results were further categorized into discordant results potentially leading to under-treatment, or potentially leading to over-treatment (<u>Supplementary Table 1</u>).^1,17^ Discordances with no clear implications for treatment were defined as “other discordances”. We assessed the adequacy of prescribed anti-TB treatment according to WHO guidelines (<u>Supplementary Table 1</u>).^1,17^ Effective drugs were defined as drugs to which *Mtb* strains had no drug resistance-conferring mutations observed in whole-genome sequences (<u>Supplementary Table 2</u>). The prescription of less than three effective drugs was defined as under-treatment, except for patients with INH- or RIF-resistant *Mtb*. In these patients, a regimen comprising less than four effective drugs was already considered as under-treatment. Over-treatment included second-line drugs given to patients for whom first-line regimens would have been adequate. <u>Supplementary Table 3</u> shows the classification of regimens in detail.

**Table 1.** Patient characteristics by resistance profiles obtained by whole-genome sequencing.

| Factor | Pan-susceptible | Any resistance | p-value* | Mono-resistance | INH | PZA | EMB | RIF | Poly-resistance | MDR | pre-/XDR | others |
| --- | --- | --- | --- | --- | --- | --- | --- | --- | --- | --- | --- | --- |
|  | 339 | 243 | ≤0.0001 | 35 | 8 | 2 | 1 | 24 | 208 | 146 | 24 | 38 |
| **Gender** |  |  |  |  |  |  |  |  |  |  |  |  |
| *Women* | 131 (39%) | 94 (39%) | ≤0.05 | 10 (29%) | 3 (38%) | 0 (0%) | 1 (100%) | 6 (25%) | 84 (40%) | 56 (38%) | 13 (54%) | 15 (39%) |
| *Men* | 208 (61%) | 149 (61%) | ≤0.01 | 25 (71%) | 5 (62%) | 2 (100%) | 0 (0%) | 18 (75%) | 124 (60%) | 90 (62%) | 11 (46%) | 23 (61%) |
| **Age** |  |  |  |  |  |  |  |  |  |  |  |  |
| *At diagnosis (IQR)* | 35 [28, 45] | 32 [25, 40] | 0.75 | 32 [25, 40] | 40 [31.25, 48.5] | 26 [24.5, 27.5] | 36 [36, 36] | 29 [24.75, 38.5] | 32 [25.75, 40.25] | 31 [25, 39] | 29.5 [24.75, 34.25] | 36 [29.25, 43.75] |
| **HIV status** |  |  |  |  |  |  |  |  |  |  |  |  |
| *HIV-negative* | 169 (50%) | 166 (68%) | 0.87 | 23 (66%) | 7 (88%) | 1 (50%) | 0 (0%) | 15 (62%) | 143 (69%) | 103 (71%) | 14 (58%) | 26 (68%) |
| *HIV-positive* | 170 (50%) | 77 (32%) | ≤0.0001 | 12 (34%) | 1 (12%) | 1 (50%) | 1 (100%) | 9 (38%) | 65 (31%) | 43 (29%) | 10 (42%) | 12 (32%) |
| **Mtb Lineage** |  |  |  |  |  |  |  |  |  |  |  |  |
| *L1* | 18 (5%) | 6 (2%) | ≤0.05 | 2 (6%) | 0 (0%) | 0 (0%) | 0 (0%) | 2 (8%) | 4 (2%) | 1 (1%) | 0 (0%) | 3 (1%) |
| *L2* | 79 (23%) | 56 (17%) | ≤0.05 | 7 (20%) | 3 (38%) | 1 (50%) | 0 (0%) | 3 (12%) | 49 (13%) | 23 (16%) | 8 (Inf%) | 18 (47%) |
| *L3* | 15 (4%) | 3 (1%) | ≤0.01 | 0 (0%) | 0 (0%) | 0 (0%) | 0 (0%) | 0 (0%) | 3 (1%) | 2 (1%) | 1 (4%) | 0 (0%) |
| *L4* | 225 (66%) | 178 (66%) | ≤0.05 | 26 (74%) | 5 (62%) | 1 (50%) | 1 (100%) | 19 (79%) | 152 (73%) | 120 (82%) | 15 (62%) | 17 (45%) |
| *L5* | 1 (<1%) | 0 (0%) | 0.32 | 0 (0%) | 0 (0%) | 0 (0%) | 0 (0%) | 0 (0%) | 0 (0%) | 0 (0%) | 0 (0%) | 0 (0%) |
| *L6* | 1 (<1%) | 0 (0%) | 0.32 | 0 (0%) | 0 (0%) | 0 (0%) | 0 (0%) | 0 (0%) | 0 (0%) | 0 (0%) | 0 (0%) | 0 (0%) |
| **Country** |  |  |  |  |  |  |  |  |  |  |  |  |
| *Côte d'Ivoire* | 46 (14%) | 48 (20%) | 0.84 | 5 (14%) | 2 (25%) | 0 (0%) | 1 (100%) | 2 (8%) | 43 (21%) | 39 (27%) | 3 (12%) | 1 (3%) |
| *Democratic Republic of the Congo* | 29 (9%) | 30 (12%) | 0.9 | 1 (3%) | 0 (0%) | 0 (0%) | 0 (0%) | 1 (4%) | 29 (14%) | 19 (13%) | 8 (33%) | 2 (5%) |
| *Kenya* | 21 (6%) | 7 (6%) | ≤0.01 | 1 (3%) | 1 (12%) | 0 (0%) | 0 (0%) | 0 (0%) | 6 (3%) | 5 (3%) | 0 (0%) | 1 (3%) |
| *Nigeria* | 19 (6%) | 34 (14%) | ≤0.05 | 6 (0%) | 0 (0%) | 0 (0%) | 0 (0%) | 6 (25%) | 28 (13%) | 20 (14%) | 4 (17%) | 4 (11%) |
| *South Africa* | 111 (33%) | 61 (25%) | ≤0.001 | 15 (43%) | 0 (0%) | 1 (50%) | 0 (0%) | 14 (58%) | 46 (22%) | 28 (19%) | 7 (29%) | 11 (29%) |
| *Peru* | 57 (17%) | 36 (15%) | ≤0.05 | 2 (6%) | 2 (25%) | 0 (0%) | 0 (0%) | 0 (0%) | 34 (16%) | 28 (19%) | 2 (8%) | 4 (11%) |
| *Thailand* | 56 (17%) | 27 (116%) | ≤0.01 | 5 (14%) | 3 (38%) | 1 (50%) | 0 (0%) | 1 (4%) | 22 (11%) | 7 (5%) | 0 (0%) | 15 (39%) |
| **History of TB** |  |  |  |  |  |  |  |  |  |  |  |  |
| *No* | 269 (79%) | 104 (43%) | ≤0.0001 | 13 (37%) | 7 (88%) | 1 (50%) | 1 (100%) | 4 (17%) | 91 (44%) | 56 (38%) | 5 (21%) | 30 (79%) |
| Yes | 70 (21%) | 139 (79%) | ≤0.0001 | 22 (63%) | 1 (12%) | 1 (50%) | 0 (0%) | 20 (83%) | 117 (56%) | 90 (62%) | 19 (79%) | 8 (21%) |
| **Treatment success** |  |  |  |  |  |  |  |  |  |  |  |  |
| *Cured* | 189 (56%) | 83 (78%) | ≤0.0001 | 10 (29%) | 3 (38%) | 0 (0%) | 1 (100%) | 6 (25%) | 73 (35%) | 45 (31%) | 8 (33%) | 20 (53%) |
| *Completed* | 59 (17%) | 46 (19%) | 0.2 | 6 (17%) | 2 (25%) | 0 (0%) | 0 (0%) | 4 (17%) | 40 (19%) | 31 (21%) | 3 (12%) | 6 (16%) |
| *Died* | 19 (6%) | 45 (19%) | ≤0.01 | 6 (17%) | 1 (12%) | 1 (50%) | 0 (0%) | 4 (17%) | 39 (19%) | 24 (16%) | 9 (38%) | 6 (16%) |
| *Treatment failure* | 11 (3%) | 10 (4%) | 0.83 | 3 (9%) | 0 (0%) | 1 (50%) | 0 (0%) | 2 (8%) | 7 (3%) | 5 (3%) | 2 (8%) | 0 (0%) |
| *Defaulted* | 20 (6%) | 22 (9%) | 0.76 | 0 (9%) | 0 (0%) | 0 (0%) | 0 (0%) | 3 (12%) | 19 (9%) | 17 (12%) | 0 (0%) | 2 (5%) |
| *Lost to follow up* | 6 (2%) | 7 (3%) | 0.78 | 2 (6%) | 0 (0%) | 0 (0%) | 0 (0%) | 2 (8%) | 5 (2%) | 5 (3%) | 0 (0%) | 0 (0%) |
| *Ongoing, unknown* | 22 (6%) | 15 (6%) | 0.25 | 3 (97%) | 2 (25%) | 0 (0%) | 0 (0%) | 1 (4%) | 12 (6%) | 9 (6%) | 0 (0%) | 3 (8%) |
| **Sputum** |  |  |  |  |  |  |  |  |  |  |  |  |
| *Positive* | 264 (80%) | 205 (85%) | ≤0.01 | 25 (71%) | 7 (88%) | 1 (50%) | 1 (100%) | 16 (67%) | 180 (87%) | 129 (88%) | 17 (74%) | 34 (92%) |
| *Negative* | 68 (20%) | 36 (15%) | ≤0.01 | 10 (29%) | 1 (12%) | 1 (50%) | 0 (0%) | 8 (33%) | 26 (13%) | 17 (12%) | 6 (26%) | 3 (8%) |

P-value\*, difference between pan-susceptible and any resistance with chi-square test. The category others includes following drug resistance: CS; EMB,RIF; ETH; INH,ETH; INH,PZA; PAS,INH,OFX,CIP,MFX,LFX; SM; SM,EMB; SM,EMB,RIF,OFX,CIP,MFX,LFX; SM,INH; SM,INH,EMB; SM,INH,ETH; SM,PZA,ETH,RIF. Abbreviations: IQR, interquartile range; *Mtb*, *Mycobacterium tuberculosis*; MDR, multi-drug-resistant; pre-XDR/XDR, pre-/ extensively-drug- resistant; WGS, whole-genome sequencing; SM, streptomycin; PAS, para-aminosalicylic-acid; INH, isoniazid; PZA, pyrazinamide, CS, cycloserine; ETH, ethionamide; EMB, ethambutol; RIF, rifampicin; OFX, ofloxacin; CIP, ciprofloxacin; MFX moxifloxacin; LFX, levofloxacin.

**Table 2:** Drug resistance results from whole-genome sequencing and local testing by diagnosis concordance and consequences for treatment.

|  | Drug resistance categories based on: |  | Diagnosis method: |  |  |  |
| --- | --- | --- | --- | --- | --- | --- |
|  | Diagnosis with WGS | Diagnosis at sites | Xpert MTB/RIF<sup>+</sup> | Culture | LPA | Combination of tests |
| **Concordant diagnosis** |  |  |  |  |  |  |
| **Total: 452 (100%)** |  |  | **242 (100%)** | **195 (100%)** | **60 (100%)** | **102 (100%)** |
| 293 (65%) | Pan-susceptible | Pan-susceptible | 196 (81%) | 139 (71%) | 49 (82%) | 53 (52%) |
| 5 (1%) | Monoresistance:<br>3 INH<br>2 RIF | Monoresistance:<br>3 INH<br>2 RIF | 0 (0%) | 4 (2%) | 1 (1.67%) | 0 (0%) |
| 138 (31%) | MDR | MDR | 45 (18.6%) | 44 (23%) | 8 (13%) | 44 (43%) |
| 9 (2%) | pre-XDR/XDR | pre-XDR/XDR | 1 (<1%) | 1 (<1%) | 2 (3%) | 5 (5%) |
| 7 (2%) | Others:<br>7 SM. | Others:<br>7 SM | 0 (0%) | 7 (4%) | 0 (0%) | 0 (0%) |
| **Discordant diagnosis** |  |  |  |  |  |  |
| **Total: 130 (100%)** |  |  | **35 (100%)** | **55 (100%)** | **9 (100%)** | **46 (100%)** |
| **Potentially leading to under-treatment** |  |  |  |  |  |  |
| **Total: 35 (100%)** |  |  | **17 (100%)** | **12 (100%)** | **1 (100%)** | **5 (100%)** |
| 0 (0%) | Pan-susceptible | - | 0 (0%) | 0 (0%) | 0 (0%) | 0 (0%) |
| 3 (9%) | Monoresistance:<br>3 INH | 3 Pan-susceptible | 2 (12%) | 1 (8%) | 0 (0%) | 0 (0%) |
| 5 (14%) | MDR | 3 Pan-susceptible<br>1 RIF<br>1 SM;EMB | 2 (12%) | 1 (8%) | 1 (100%) | 1 (20%) |
| 15 (43%) | pre-XDR/XDR | MDR | 5 (29%) | 6 (50%) | 0 (0%) | 4 (80%) |
| 12 (34%) | Others:<br>10 INH;ETH<br>1 SM;INH;ETH<br>1 SM;INH | Pan-susceptible<br>Pan-susceptible<br>Pan-susceptible | 8 (47%) | 4 (33%) | 0 (0%) | 0 (0%) |
| **Potentially leading to over-treatment** |  |  |  |  |  |  |
| **Total: 31 (100%)** |  |  | **3 (100%)** | **12 (100%)** | **2 (100%)** | **14 (50%)** |
| 23 (74%) | Pan-susceptible | 1 INH<br>18 MDR<br>4 RIF | 3 (100%) | 9 (75%) | 2 (100%) | 9 (64%) |
| 2 (6%) | Monoresistance:<br>2 INH | MDR | 0 (0%) | 2 (16.67%) | 0 (0%) | 0 (0%) |
| 3 (10%) | MDR | pre-XDR/XDR | 0 (0%) | 0 (0%) | 0 (0%) | 3 (21%) |
| 0 (0%) | pre-XDR/XDR | - | 0 (0%) | 0 (0%) | 0 (0%) | 0 (0%) |
| 3 (10%) | Others:<br>1 INH;PZA<br>1 SM;INH;EMB<br>1 INH;ETH | MDR<br>MDR<br>MDR | 0 (0%) | 1 (8%) | 0 (0%) | 2 (14%) |
| **Other discordance in diagnosis** |  |  |  |  |  |  |
| **Total: 64 (100%)** |  |  | **15 (100%)** | **31 (100%)** | **6 (100%)** | **27 (100%)** |
| 23 (36%) | Pan-susceptible | 20 EMB<br>1 Monoresistance*<br>1 SM | 2 (13%) | 20 (65%) | 0 (0%) | 1 (4%) |
| 25 (39%) | Monoresistance:<br>20 RIF<br>2 PZA<br>1 EMB | MDR<br>Pan-susceptible<br>Pan-susceptible | 6 (40%) | 1 (3%) | 1 (17%) | 20 (74%) |
| 0 (0%) | MDR | - | 0 (0%) | 0 (0%) | 0 (0%) | 0 (0%) |
| 0 (0%) | pre-XDR/XDR | - | 0 (0%) | 0 (0%) | 0 (0%) | 0 (0%) |
| 16 (25%) | Others:<br>5 ETH<br>1 CS.<br>1 SM.<br>3 INH;ETH<br>1 SM;EMB<br>1 SM;EMB;RIF;OFX;CIP;MFX;LFX<br>1 SM.<br>1 PAS;INH;OFX;CIP;MFX;LFX<br>1 SM;PZA;ETH;RIF<br>1 EMB;RIF | Pan-susceptible<br>Pan-susceptible<br>SM;EMB<br>INH<br>SM<br>MDR<br>Pan-susceptible<br>INH<br>MDR<br>MDR | 7 (47%) | 10 (32%) |  | 6 (22%) |

Abbreviations: vs, versus; WGS, whole-genome sequencing; *Mtb*, Mycobacterium tuberculosis; DST, drug susceptibility test; LPA, line probe assay; MDR, multi-drug-resistant; pre-XDR/XDR, pre-/extensively drug-resistant; WHO, World Health Organization; SM, streptomycin; PAS, para-aminosalicylic-acid; INH, isoniazid; PZA, pyrazinamide; CS, cycloserine; ETH, ethionamide; EMB, ethambutol; RIF rifampicin; OFX, ofloxacin; CIP, ciprofloxacin; MFX moxifloxacin; LFX, levofloxacin. \*exact Monoresistance is not known; \*RIF resistance diagnosed with Xpert MTB/RIF was classified as MDR.

**Table 3:** Mortality by concordance of local diagnosis and whole-genome sequencing.

|  | Total | Concordant with diagnosis at sites | Discordant with diagnosis at sites |  |  |  |
| --- | --- | --- | --- | --- | --- | --- |
|  |  |  | Any discordance | Potentially leading to under-treatment | Potentially leading to over-treatment | Other discordance |
| **Resistance based on WGS:** | 63/530 (12%) | 44/409 (11%) | 19/121 (16%) | 8/30 (27%) | 1/28 (4%) | 10/63 (16%) |
| Pan-susceptible | 18/310 (6%) | 16/267 (6%) | 2/43 (5%) | 0/0 | 0/20 (0%) | 2/23 (9%) |
| Any resistance | 45/220 (20%) | 28/142 (20%) | 17/78 (22%) | 8/30 (27%) | 1/8 (12%) | 8/40 (20%) |
| **Mono-resistance:** | 6/32 (19%) | 2/4 (50%) | 4/28 (14%) | 0/1 (0%) | 0/2 (0%) | 4/25 (16%) |
| INH | 1/6 (17%) | 1/3 (33%) | 0/3 (0%) | 0/1 (0%) | 0/2 (0%) | 0/0 |
| PZA | 1/2 (50%) | 0/0 | 1/2 (50%) | 0/0 | 0/0 | 1/2 (50%) |
| EMB | 0/1 (0%) | 0/0 | 0/1 (0%) | 0/0 | 0/0 | 0/1 (0%) |
| RIF | 4/23 (17%) | 1/1 (100%) | 3/22 (14%) | 0/0 | 0/0 | 3/22 (14%) |
| **Poly-resistance:** | 39/188 (21%) | 26/138 (19%) | 13/50 (26%) | 8/29 (28%) | 1/6 (17%) | 4/15 (27%) |
| MDR | 24/131 (18%) | 23/123 (19%) | 1/8 (12%) | 1/5 (20%) | 0/3 (0%) | 0/0 |
| pre-XDR/XDR | 9/23 (39%) | 2/8 (25%) | 7/15 (47%) | 7/15 (47%) | 0/0 | 0/0 |
| Others | 6/34 (18%) | 1/7 (14%) | 5/27 (19%) | 0/9 (0%) | 1/3 (33%) | 4/15 (27%) |

Analysis based on 530 patients with complete data. The estimated mortality is calculated by comparing the deaths compared to the total of the category. Abbreviations: WGS, whole-genome sequencing; MDR, multi-drug-resistant; pre-XDR/XDR, pre-/extensively drug-resistant; Others: CS; EMB,RIF; ETH; INH,ETH; INH,PZA; PAS,INH,OFX,CIP,MFX,LFX; SM; SM,EMB; SM,EMB,RIF,OFX,CIP,MFX,LFX; SM,INH; SM,INH,EMB; SM,INH,ETH; SM,PZA,ETH,RIF; SM, streptomycin; PAS, para-aminosalicylic-acid; INH, isoniazid; PZA, pyrazinamide, CS cycloserine; ETH, ethionamide; EMB, ethambutol; RIF, rifampicin; OFX, ofloxacin; CIP, ciprofloxacin; MFX, moxifloxacin; LFX, levofloxacin.

### Statistical methods

We used descriptive statistics to assess patient characteristics by levels of drug resistance based on WGS. We calculated the agreement between the results of local and WGS drug resistance results using Cohen’s Kappa statistic (in R package “psych”).^18,19^ We compared the following drug resistance categories: pan-susceptible TB, monoresistant TB (any monoresistance), MDR-TB, pre-XDR/XDR-TB, any INH-resistant TB (including INH-monoresistant TB, MDR-TB and pre-XDR/XDR-TB), any RIF-resistant TB (including RIF-monoresistant TB, MDR-TB and pre-XDR/XDR-TB). We examined the determinants of mortality in univariable and multivariable logistic regression models. Patients with missing data on treatment regimen, treatment outcome, ongoing treatment, and sputum microscopy, respectively, were excluded from the analysis of mortality.

Four separate logistic regression models were performed to assess the impact on mortality: 1) the impact of any drug resistance on mortality, 2) the impact of different types of drug resistance on mortality, 3) the impact of discordant diagnoses on mortality, and 4) the impact of treatment adequacy (according to WHO guidelines) on mortality. Logistic regression models were adjusted for sex, age, HIV-status, history of TB, sputum positivity, and *Mtb* lineage (<u>Supplementary Table 4</u>). All analyses were performed in R v3.6.1 or Python v3.7.6.^18,20^ We conducted two sensitivity analyses. First, we repeated all logistic regression analyses after restricting the data to drug resistances that could be diagnosed with the locally available tests. We thus excluded drug resistances that were missed due to unavailable testing methods. Second, we examined the impact of different variant frequency cut-offs: > 0% and 100%.

### Ethics

The Cantonal Ethics Committee in Bern, Switzerland, and local institutional review boards approved the study. Written informed consent was obtained at all sites, except in South Africa, where consent was not required for archived samples.

## Results

### Study population

Of the 634 patients included in our previous analysis,^6^ we failed to sequence 52 (8.2%) *Mtb* isolates for technical reasons (<u>Supplementary Figure 1</u>). We thus included 582 TB patients, 406 (70%) from Africa, 83 (14%) from Asia, and 93 (16%) from Latin America. A total of 172 patients (30%) came from South Africa, 94 (16%) from Côte d’Ivoire, 59 (10%) from the Democratic Republic of the Congo, 28 (5%) from Kenya, 53 (9%) from Nigeria, 93 (16%) from Peru, and 83 (14%) from Thailand (Table 1). The median age was 32 years (interquartile range: 27-43 years), 225 (39%) were female, and 247 (42%) were HIV-positive. Six *Mtb* lineages were represented: 24 (4%) L1, 135 (23%) L2, 18 (3%) L3, 403 (69%) L4, one (<1%) L5, and one (<1%) L6. Based on WGS, 339 (58%) isolates were pan-susceptible, 35 (6%) monoresistant, including 24 (4%) RIF, 8 (1%) INH, 2 (<1%) PZA and 1 (<1%) EMB mono-resistant isolates. There were 146 (25%) MDR, and 24 (4%) pre-XDR/XDR isolates (Table 1; Figure 1). Among the 24 patients with pre-XDR/XDR, 9/24 (38%) had resistance to FQLs, 6 (25%) to INJ, and 9 (38%) to both.

**Figure 1.**
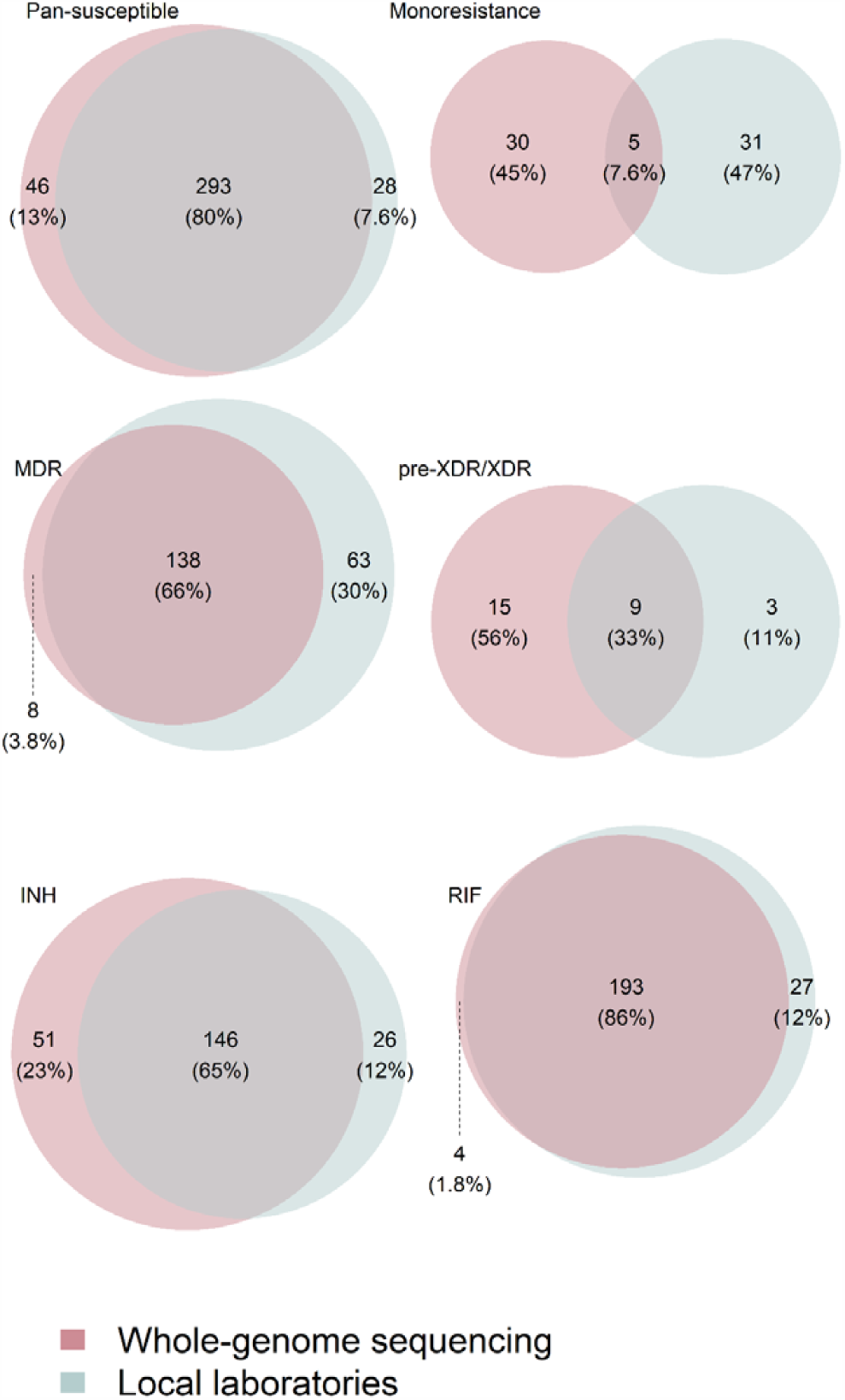
Distribution of diagnosed drug resistance between whole-genome sequencing and local DST. The categories include pan-susceptible TB, monoresistant TB (any monoresistance), MDR-TB, pre-XDR/XDR-TB, any INH-resistant TB, any RIF-resistant TB. Abbreviations: WGS, whole-genome sequencing; DST, drug susceptibility testing; MDR, multi-drug-resistant; pre-XDR/XDR, pre- /extensively drug-resistant; INH, isoniazid; RIF rifampicin.

### Local results of DST

Local DST results were based on the molecular Xpert MTB/RIF test system, line probe assays (LPA), and culture-based phenotypic tests or a combination of these methods (Table 2). Among the 582 *Mtb* isolates, 130/582 (22%) had discordant drug resistance results that potentially led to inadequate treatments in 66/582 (11%) of TB patients (Table 2). In 35/50 (70%) patients, under-diagnosis potentially led to under-treatment, and in 31/80 (39%), over-diagnosis potentially led to over-treatment (Table 2). For six patients, we had no treatment information. Of 576 patients with known treatment, we observed that overall, 86/576 (15%) patients received inadequate treatment according to WGS results and WHO treatment guidelines: 67/576 (12%) patients were under-treated, and 19 (3%) were over-treated. Consequently, 490/576 (85%) were adequately treated.

The agreement between local DST and WGS (as assessed by Cohen’s kappa) were 0.74 (95% CI 0.69-0.8) for pan-susceptible, 0.09 (-0.03-0.2) for mono-resistant, 0.71 (0.65-0.77) for MDR, and 0.49 (0.28-0.69) for pre-XDR/XDR-TB cases (Figure 1). Agreement of local DST and WGS for RIF resistance were 0.88 (0.84-0.92) and 0.69 (0.63-0.76) for INH resistance (Figure 1). RIF resistance was, in contrast to other drug resistance, more frequently diagnosed with local DST than with WGS (Figure 1). Only three sites tested other drugs than RIF and INH. Two sites tested for SM, two for FQLs, and two for INJ. One site tested for PZA and one site for EMB. Resistance to PAS, CS, EMB, LZD, BDQ, CFZ, and DLM was not tested at any site. Resistance to the latter three was also not diagnosed with WGS using a threshold of ≥ 90% variant frequency (<u>Supplementary Table 5</u>).

### Mortality

We excluded 52/582 (9%) patients due to missing data (Table 1; <u>Supplementary Figure 1</u>). The included patients’ median age was 34 years (interquartile range: 27-43 years), 207 (39%) were female, and 218 (41%) were HIV-positive. Based on WGS, the *Mtb* isolates of 310 (58%) patients were pan-susceptible, 32 (6%) mono-resistant, 131 (25%) MDR and 23 (4%) pre-XDR/XDR. Among the 530 isolates, 121 (23%) had a discordant diagnosis. For 30/44 (68%) patients, the under-diagnosis of drug resistance potentially led to under-treatment, and for 28/77 (36%) the over-diagnosis potentially led to over-treatment.

During treatment, 63/530 (12%) patients died (Table 3). Patients with pan-susceptible *Mtb* had a mortality of 18/310 (6%), patients with monoresistant TB had a mortality of 6/32 (19%), patients with MDR-TB had a mortality of 24/131 (18%), and patients with pre-XDR/XDR-TB had a mortality of 9/23 (39%) (Figure 2A). Overall, mortality ranged from 16/267 (6%) among patients with pan-susceptible strains and concordant diagnosis to 7/15 (47%) among patients with pre-XDR/XDR-TB and a discordant diagnosis potentially leading to under-treatment (Table 3). In patients with a discordant diagnosis potentially leading to under-treatment, mortality was 8/30 (27%), and in patients with discordant diagnosis potentially leading to over-treatment, it was 1/28 (4%) (Figure 2B). Mortality ranged from 17/293 (6%) in patients with pan-susceptible TB treated according to WHO guidelines to 19/60 (32%) in under-treated patients and 1/17 (6%) in patients who were over-treated (Figure 2C).

**Figure 2.**
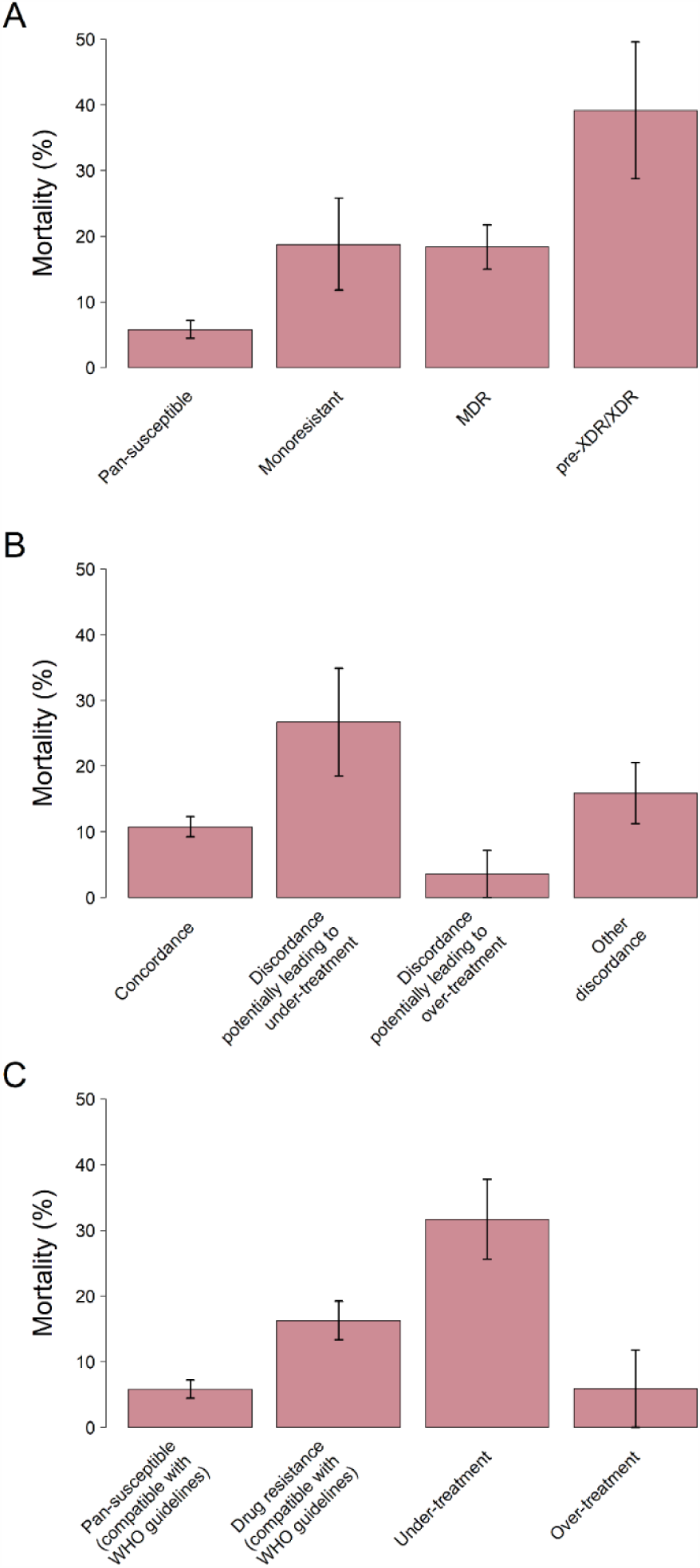
Mortality according to drug resistance, concordance of diagnosis, and treatment adequacy (according to WHO guidelines, see also supplementary tables 1 and 2). Error bars are standard errors. Analysis based on 530 patients with complete data. The estimated mortality is calculated by comparing the deaths compared to the total amount of the category. Abbreviations: WHO, World Health Organization; MDR, multi-drug-resistant; pre-XDR/XDR, pre-extensively and extensively drug-resistant.

In the multivariable logistic regression adjusted for sex, age, HIV status, history of TB, sputum microscopy, and TB lineage, resistance to any of the anti-TB drugs was associated with higher mortality (Figure 3; <u>Supplementary Table 4</u>): the adjusted odds ratio (aOR) was 5.71 (95% CI 3.02-11.25). The association with mortality became stronger with higher degree of drug resistance. Compared to pan-susceptible TB, the odds ratios for mono-resistant, MDR-, and pre-XDR/XDR TB were 5.40 (95% CI 1.73-15.49), 5.76 (95% CI 2.71-12.54), and 22.07 (95% CI 7.21-68.27), respectively (Figure 3). The aOR for mortality during TB treatment was 3.82 (95% CI 1.47-9.25) in patients with a diagnosis potentially leading to under-treatment, and 0.28 (95% CI 0.02-1.39) in the case of a diagnosis potentially leading to over-treatment, compared to patients with concordant treatment (Figure 3). Overall, 77/530 (15%) patients received inadequate treatment based on WGS drug resistance results and WHO guidelines (<u>Supplementary Table 3</u>): 60/530 (11%) patients were under-treated, and 17/530 (3%) were over-treated. The odds ratio for mortality for under-treatment was 4.82 (95% CI 2.43-9.44), and for over-treatment, it was 0.52 (95% CI 0.03-2.73), compared to patients receiving adequate treatment (Figure 3).

**Figure 3.**
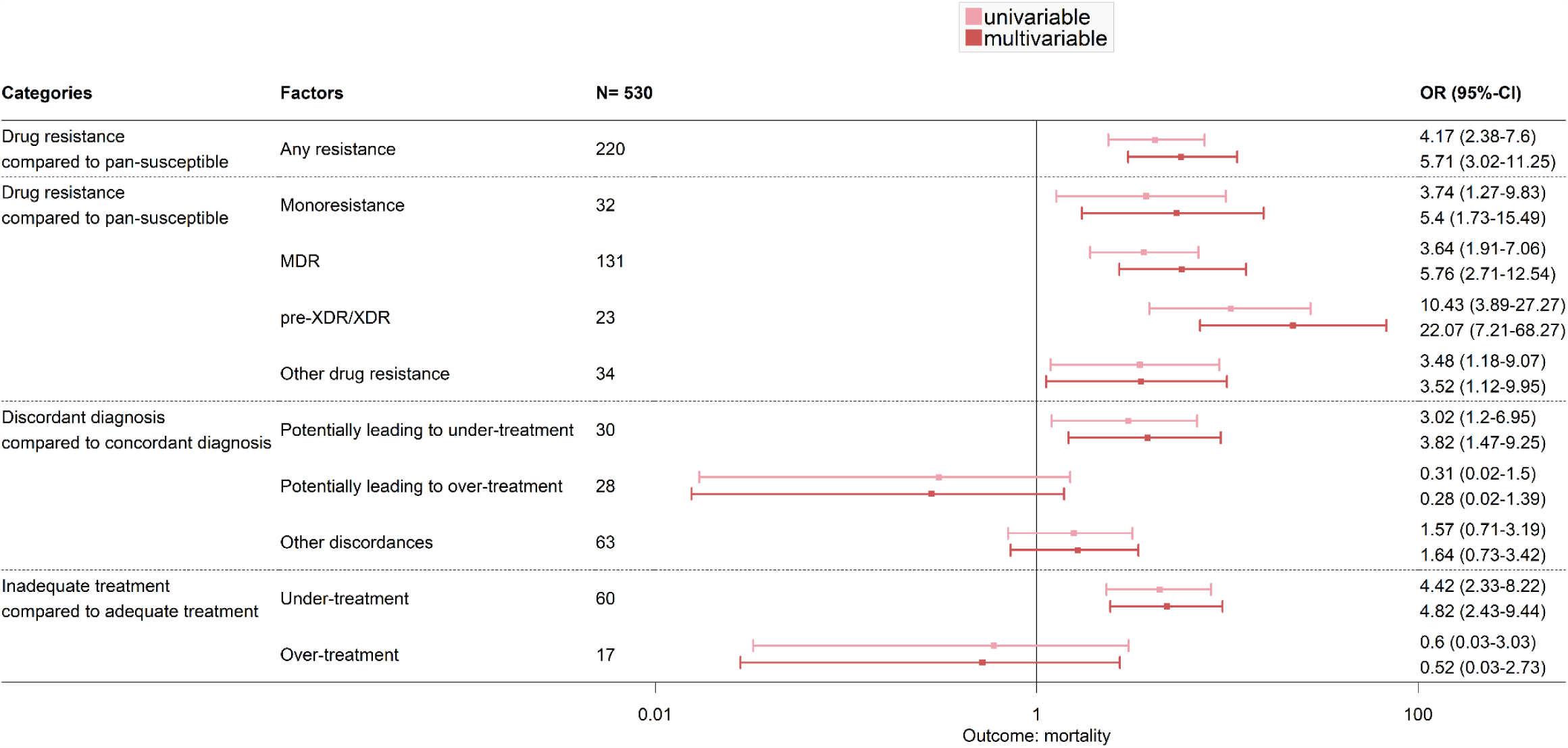
Results from four logistic regression models (adjusted for sex, age, HIV status, history of TB, sputum microscopy, and TB lineage) to assess 1) the impact of any drug resistance on mortality, 2) the impact of drug resistance categories on mortality, 3) the impact of diagnosis discordance on mortality, 4) the impact of treatment adequacy (according to WHO guidelines) on mortality. Abbreviations: OR, odds ratio; CI, confidence interval; N, number; *Mtb, Mycobacterium tuberculosis*; WHO, world health organisation; MDR, multi-drug-resistant; pre-XDR/XDR, pre-extensively and extensively drug-resistant.

The sensitivity analysis using different variant frequency cut-offs (> 0% and 100%) produced similar results (<u>Supplementary Figure 2 and 3</u>). When restricting the analysis to drug resistances which could be diagnosed at sites, again similar results were obtained (<u>Supplementary Figure 4</u>).

## Discussion

Drug-resistant TB is a significant global health problem. In this multicentre cohort study of 582 TB patients, we compared drug resistance predicted by WGS with the results from local DST in eight high-burden TB countries. We examined mortality by drug resistance predicted by WGS, and by concordance or discordance with local diagnosis and treatment. We found that the diagnosis was discordant between local drug resistance results and WGS in about one in five patients. The agreement between local and centralized WGS was the highest for RIF and INH but low for other drugs. Of note, resistance to SM, PAS, PZA, CS, ETH, EMB, FQLs, and INJ was rarely investigated locally. Mortality during treatment ranged from 6% among patients with pan-susceptible strains and concordant results between WGS and local drug resistance testing to 47% among patients with pre-XDR/XDR TB and discordant results.

To our knowledge, this is the first study to compare the results from DST in real-world settings in high-burden countries with WGS and to examine the impact of discordant resistance results on mortality. In a previous study of this cohort, we compared the results from local DST with standardized liquid culture on six drugs performed at the Swiss National Center for Mycobacteria.^6^ In the present study, we used a well-established bioinformatics pipeline and its corresponding database to analyse the WGS data.^7^ The analysis covered 19 drugs, including SM, KM, PZA, ETH, EMB, LFX, and newer drugs. Specifically, we were able to detect more single drug resistance with WGS compared to phenotypic DST.

Rapid and accurate diagnosis, prompt and appropriate treatment, and the control of airborne infection are key strategies to prevent drug-resistant TB.^21^ WGS has the potential to predict resistance profiles for most anti-TB drugs without the need for time-consuming phenotypic DST.^1,7,8,16^ WGS allows effective individualised treatment and thus reduces the risk of propagating drug resistance. Ineffective treatment may lead to the acquisition of additional drug resistance and increased transmission of drug-resistant strains.^21^ These considerations support the use of WGS to replace the current DST methods, which cover only a limited number of drugs. Culture-based DST is challenging for several drugs, for example, PZA, ETH, and EMB, due to poor drug solubility.^8,22^ The accurate diagnosis of pertinent drug resistance is essential. For example, PZA is critical to shortening TB therapy, and resistance is related to a worse outcome.^21^ Yet, PZA resistance testing is often unavailable. Only one site could test PZA resistance. Routine DST at sites focused mainly on the identification of RIF and INH resistance, used to defined MDR-TB, leaving the efficacy of other drugs unanswered. Furthermore, INH monoresistance would typically be missed when DST relies on the use of Xpert MTB/RIF, which could lead to the under-treatment of Xpert-negative individuals. In contrast to other molecular and phenotypic DST methods, WGS simultaneously detects all known drug resistance-conferring mutations. The wider range of drug resistance captured by WGS explains some of the discordant results found in this study. However, discordant results potentially leading to inadequate treatment were mainly due to drug resistance that was undiagnosed, despite the availability of tests at sites, rather than to a wider range of drug resistances captured by WGS.

The detection of drug resistance is influenced by the type of sample collected, and the methods used for culturing, DNA extraction and sequencing, and the pipeline used to analyse the sequences.^23^ The latter was determined by a ≥ 90% variant frequency cut-off and the robustness of the TBprofiler pipeline and its coverage of all relevant resistance-conferring mutations. Our sensitivity analysis showed that the cut-off for variant frequency had little impact on results. For new drugs, most resistance-conferring mutations are unknown at the time of introduction, and relevant drug resistance mechanisms become apparent only later on. Discrepancies in results between local DST and WGS might be explained by mixed infections, heteroresistance, minority resistant populations, or methodological differences,^23-25^ and lead to uncertainties in treatment decisions.^26^ Of note, over-treatment did not increase mortality, but the analysis was based on few patients and should be interpreted with caution. Anti-TB drugs can have serious side effects, which may lead to treatment interruption and failure, or acquired drug resistance. Over-treatment should be avoided from both a medical and cost perspective.

Our study has several limitations. We sampled eligible patients within strata defined by drug resistance and HIV infection and therefore, could not estimate the incidence or prevalence of drug-resistant TB in HIV-positive or HIV-negative patients. We could also not evaluate differences in drug resistance between *Mtb* lineages, because the sample size was small for several lineages. Further, we sequenced *Mtb* strains before treatment and could thus not diagnose potential acquired drug resistance, which may influence treatment outcome. Finally, we started this study in 2013. Since then, TB treatment guidelines changed. In 2013, WHO published an interim policy guideline on BDQ and in 2014 on DLM in the treatment of MDR-TB.^27,28^ In our study, patients were rarely treated with new drugs. Consequently, our results may not reflect the situation in 2020. However, our study does show that treatment strategies guided by comprehensive drug resistance data are likely to save lives.

Our results support WHO’s call for an accurate point-of-care test based on WGS that can be performed directly from sputum samples.^29^ Such tests would allow rapid diagnosis and efficient, individual-based treatment of drug-resistant TB. Test systems performing WGS on sputum samples and bioinformatics pipelines are currently in development. High-burden countries should consider building central, high-throughput sequencing capacities. The establishment of a trustworthy, widely accepted drug resistance database similar to the Stanford HIV drug resistance database will be essential in this context.^30^ Finally, we support the call for clinical trials evaluating the efficacy, safety, tolerability of new TB treatment and DST strategies for drug-resistant TB.^21^ Especially, the role of new drugs like BDQ, DLM, and pretomanid in regimens with fewer, more effective and safer drugs needs to be evaluated.^21^ Future studies should also examine treatment duration and adherence.^21^ The duration of the intensive and continuation phases of TB treatment and treatment adherence are crucial for efficient therapy.

In conclusion, our study shows that both the accuracy of DST in routine care and the access to testing for resistance for several essential drugs is limited in high-burden TB countries and that this leads to inadequate treatment and contributes to higher mortality. Our results support the role of WGS to improve the management of drug-resistant TB in high-burden TB settings.

## Data Availability

Genome data from patients' Mycobacterium tuberculosis strains included in this analysis have been submitted to the NCBI (PRJNA300846; Supplementary Table 6).

## Data availability

WGS data from patients *Mtb* strains included in this analysis have been submitted to the NCBI (PRJNA300846; <u>Supplementary Table 6</u>).

## Acknowledgement

We thank all sites that participated, patients whose data were used in this study. We are also grateful to the Tuberculosis Working Group of IeDEA for helpful discussions. This research was supported by the Swiss National Foundation (project grant numbers 153442, 310030_188888, IZRJZ3_164171, IZLSZ3_170834, CRSII5_177163 and special project grant 17481). The International Epidemiology Databases to Evaluate AIDS (IeDEA) is supported by the US National Institutes of Health, National Institute of Allergy and Infectious Diseases, the Eunice Kennedy Shriver National Institute of Child Health and Human Development, the National Cancer Institute, the National Institute of Mental Health, the National Institute on Drug Abuse, the National Heart, Lung, and Blood Institute, the National Institute on Alcohol Abuse and Alcoholism, the National Institute of Diabetes and Digestive and Kidney Diseases, the Fogarty International Center, and the National Library of Medicine: Asia-Pacific, U01AI069907; CCASAnet, U01AI069923; Central Africa, U01AI096299; East Africa, U01AI069911; NA-ACCORD, U01AI069918; Southern Africa, U01AI069924; West Africa, U01AI069919. RJW receives support from the Francis Crick Institute, which is funded by UKRI, CRUK (FC0010218), and Wellcome (104803, 203135). This work is solely the responsibility of the authors and does not necessarily represent the official views of any of the institutions mentioned above. Calculations were performed on UBELIX (http://www.id.unibe.ch/hpc), the HPC cluster at the University of Bern.

## Notes

### Competing Interest Statement

MLR, KZ, MB, CL, SB, MR, VS, RH, PS, AA, AGA, OM, JC, EJC, HC, RH, LF, SG, and ME have nothing to disclose. RJW reports grants from Wellcome, grants from European and Developing Countries Clinical Trials Partnership, grants from UK Research and innovation, grants from Cancer Research UK, grants from National Institutes of Health, during the conduct of the study. ECB reports personal fees from AID Diagnostika, personal fees from COPAN, outside the submitted work. MY reports grants from US National institute of Health, during the conduct of the study.

